# Effect of Finger Millet-Based Supplementation on Anthropometry and Body Composition Among Moderately Acute Malnourished Under-5 Children in Telangana, India: A Randomized, Community-Based Trial

**DOI:** 10.1101/2025.11.21.25340731

**Authors:** B R Nikhita, Sakshi Rai, Parth Sarin, Sahil Bipin Kumar Suthar, Hemant Mahajan, Aruna V. Reddy, G Vijayalakshmi, Santosh Kumar Banjara, Ananthan Rajendran, R Naveen Kumar, Sudhakar Ajmera, Sourav Sengupta, Devaraj J. Parasannanavar

**Author notes:** Corresponding Author: Correspondence: Devaraj J. Parasannanavar, Scientist D Maternal and Child Health Nutrition Division, ICMR-National Institute of Nutrition, Hyderabad., Email ID, Contact: 8074928620.

## Abstract

**Background:** Moderate acute malnutrition (MAM) remains a major health challenge in India despite ongoing efforts of government nutrition programs. Finger millet (Ragi), a nutrient-dense, climate-resilient crop rich in calcium, fiber, and bioactive compounds, offers potential advantages over conventional cereal-based ready-to-use therapeutic foods (RUTF). This trial evaluated the effects of finger millet-based supplement on anthropometry, body composition, dietary adequacy among MAM children aged 18-59 months.

**Material & methods:** In this open-label, randomized community-based trial, 221 MAM children were allocated to receive either a finger millet-with-dates supplement (FMD) or the standard wheat/rice-based Balamrutham plus with dates (BMD) for 8 weeks in Anganwadi centers in Hyderabad, India. Anthropometry, body composition (InBody S10), dietary intake (three-day recall), biomarkers, and compliance were assessed at baseline and endpoint, with follow-ups at days 100 and 160. Growth trends were evaluated using repeated-measures ANOVA.

**Results:** 136 children completed the intervention (FMD: 71; BMD: 65); compliance exceeded 80%. By day 40, both groups showed significant gains in weight (6.3-6.4%), height (2.4%), and MUAC (3.5-6.2%), which continued through day 160. The FMD group showed greater improvements in mineral mass, bone mineral content, and skeletal muscle mass, whereas the BMD group demonstrated higher increases in protein mass and fat-free mass. Dietary recall revealed markedly low micronutrient adequacy among MAM children. No major adverse events were reported.

**Conclusion:** Finger millet-based supplementation achieved comparable or superior improvements to cereal-based RUTF, supporting its integration into ICDS and POSHAN 2.0 as a sustainable approach to MAM management.

## Introduction

Children’s nutritional status is a key indicator of a nation’s economic development. An essential part in global health care is attaining Optimal Nutrition, especially in the management of Childhood malnutrition and its severe consequences (Choge et al., 2020). Malnutrition denotes a mismatch between body’s nutrient intake and requirements. It manifests as dietary excesses or imbalances and represents a persistent public health challenge that garners worldwide attention. It is closely intertwined with multiple sustainable developmental goals (SDGs) adopted by United Nations 2015 stating malnutrition reduction as one of their goals (SDG 2.2). The World Health Organization (WHO) categorizes malnutrition into three types: i) Undernutrition (Stunting, wasting and underweight) seen mostly in children, refers to inadequate consumption, poor absorption and excessive nutrient loss ii) Micronutrient related malnutrition and iii) Overnutrition –overweight/ obesity with diet related non-communicable diseases (Chakravorty S et al.,2023).

Undernutrition in children under five is a universal health problem and a major contributor to higher rates of morbidity and mortality. It also hampers cognitive skills and physical growth. The Global nutrition report (2021) states that about 7.7% of children were severely wasted, 19.3% were wasted and 35.5% were stunted. In India, Undernutrition remains a leading cause of child mortality despite its rapid economic surge. The 2015-16 National Survey indicated that 35.7% of under 5 children were underweight, 28.5% suffered from wasting and 38.4% experienced stunting. The associated risk factors include individual biological and behavioral traits, food insecurity, socio-economic status, low dietary intake, poor dietary diversity, inadequate maternal nutrition, infections, and insufficient environmental hygiene and sanitation. Although children need abundant essential nutrients for their overall growth and development, they often fall short of meeting these requirements due to above risk factors. Current studies indicate that interventions aimed at preventing undernutrition should prioritize this vulnerable group, as this is a critical factor for India in reaching the global 2030 targets set by the WHO to reduce malnutrition worldwide (Shah et al., 2024).

In India, numerous government initiatives, including Integrated Child Development Services (ICDS), National Health Mission, Poshan 2.0, and Mid-Day Meal schemes, have been launched to enhance child health. These programs promote nutritional education and offer Ready-to-Use Therapeutic Foods (RUTF), typically cereal-based (calorie dense formulas) to support child’s growth. In partnership with Indian Council of Medical Research-National Institute of Nutrition (ICMR-NIN), Telangana developed Balamrutham+, a nutrient-rich RUTF, to manage Moderate and severe acute malnutrition (MAM/SAM) in children aged 6-59 months. It is given based on the child’s weight, offering 75kcal/kg and 125kcal/kg for MAM and SAM cases respectively. Feeding is continued till the child attains healthy levels for Weight-for-height (WHZ) and Weight for Age (WAZ) scores. The main composition of the product is wheat flour, rice flakes, groundnut, Bengal gram, skimmed milk powder, sugar and oil which follows all recommended WHO guidelines for complementary foods for undernourished children (Kumar et al.,2020). However, despite these ongoing efforts, the issue of undernutrition among children remains a significant challenge.

Millet-based interventions have shown positive results in terms of child’s overall growth and improvement including body composition parameters and hemoglobin status due to their rich nutrient profile, making them a valuable addition to their diets (Anitha S et al., 2021). Millets, known as “Nutri-Cereals,” have recently gained attention for their role in improving nutrition, supporting environmental sustainability, and enhancing food security. These resilient crops, which thrive in poor soils and tough climates, are divided into major millets like Finger millet (Ragi), Pearl millet (Bajra), Sorghum (Jowar), and minor millets such as Foxtail, Barnyard, Kodo, Proso, and Little millet. These are the vital food crops in developing countries, recognized for their high energy and nutritional value. They promote overall health by preventing and managing conditions such as obesity, diabetes, and cardiovascular issues (Sarita and Singh, 2016). India is the leading producer of millets, with Finger millet (Ragi), particularly noted for its impressive calcium content (364 mg per 100 grams), well balanced amino acids, iron, polyphenols and other nutrients. Ragi is rich in dietary fiber that consists of 2-3% of soluble fiber such as beta glucan, arabinoxylan and 19-20% of insoluble fiber namely cellulose, hemicellulose and lignin which proves to a potent prebiotic source (Boda et al., 2025; Singh V et al.,2022). On contrary, finger millet is also a hub of antinutrients such as tannins, phytates and saponins etc. which can be reduced through appropriate processing methods like germination which is an easier and flexible option (Shobana et al., 2013; Anitha S et al., 2022).

In 2023, Indian government declared it as the International year of Millets. The main purpose was to launch innovative schemes that help millet cultivation and production as well as promoting its health benefits. States like Tamil Nadu, Odisha, Chhattisgarh, Madhya Pradesh and Telangana have begun incorporating millets, especially ragi into the daily meals provided by state-run schools, promoting healthier nutrition (NITI Aayog, 2023). POSHAN 2.0 scheme is one of the government initiatives that works mainly to combat malnutrition in children, adolescent girls, pregnant women, and lactating mothers. It focuses on maternal nutrition, child feeding norms, and traditional practices like using millets in meals, with an emphasis on community outreach and innovations in nutrition support. The use of Finger Millet (Ragi), to substitute for wheat/rice in complementary feeding programs in Telangana can be a key initiative to enhance nutrition for the malnourished children (Nayak B, 2023).

In our study, the Finger millet-based RUTF and wheat/rice based Balamrutham plus were supplemented among MAM children at AWCs. Date powder was used as a replacement of sugar in the existing product. Using date powder as a natural sweetener supports children’s health, while reducing the risk of issues like obesity, dental caries, and inflammation caused by excess added sugar in most complementary foods (Almuziree RSA, 2023). With a low glycaemic index, dates provide more nutrients compared to cane sugar, including fibre, iron, potassium, zinc, and copper. 100g of cane sugar has 387 kcal and no fibre, the same amount of dates offer 282 kcal, 8g of fibre, along with superior micronutrient content. In comparison to the existing SSFP wheat and rice-based product, this study aims to understand how finger millet-based diet supplementation affects the body measurements, composition, and micronutrient status in moderately acute malnourished children aged 2-5. This will be assessed through a variety of indicators, including anthropometric measurements, body composition (including fat mass, fat-free mass, body water etc.,) analysis, biochemical parameters, and clinical manifestations. Early initiation of a safe, reliable, and sustainable calcium-rich diet, especially from plant-based traditional crops, represents a promising approach to treating malnutrition. Millets, like ragi, were once commonly used in everyday diets, especially as weaning food for children. However, this tradition has faded in modern times for various reasons. Both Finger millet and date powder are micronutrient rich which can further address hidden hunger among children. This research will serve as a proof of concept for a larger randomized clinical trial that includes the use of millets to address undernutrition in children.

## Methods

### Study Design and Participants

An open-labelled, randomized controlled intervention trial was conducted across Anganwadi Centres (AWCs) in and around ICMR–NIN, Hyderabad, Telangana, India. Children with Moderate Acute Malnutrition (MAM) were randomly allocated to receive either a finger millet–based supplement or a wheat-based supplement. A ready-to-eat portion of 90–150 g/day (equivalent to approximately 75 kcal/kg body weight/day) was provided as *ladoo, idly, pancake, or porridge*, administered as morning and evening snacks for ≥5 days per week over 8 weeks. Anganwadi workers and caregivers were trained to maintain each child’s routine supplementary intake throughout the study.

Anthropometry (weight, height, MUAC), body composition (InBody S10), and venous blood samples were obtained at baseline (day 0) and endpoint (day 40). Supplementation adherence and any adverse events were monitored daily; illnesses such as fever, cough, allergies, or diarrhoea were managed by study clinicians. Nutritional recovery was defined as achieving a WHZ > –1.

### Sample Size

At the beginning of the study, the sample size was calculated using data from a small pilot conducted in nearby Anganwadi Centres. The pilot suggested that children with moderate acute malnutrition gained approximately 100 g per month, whereas healthy children gained 200–300 g per month. Evidence from earlier millet-based interventions indicated about a 25% improvement in weight gain with finger millet supplementation (Anitha et al., 2021). Based on these findings, the expected average weight gain over 8 weeks was assumed to be approximately 4 g/kg/day in the finger-millet group and 3 g/kg/day in the existing cereal-based group.

Using a multiple-comparison framework with Bonferroni correction (adjusted α = 0.016), and assuming a mean difference of 0.9 units with an SD of 1.5, the required sample was calculated in Stata 15. For 80% power and allowing for 5% attrition, the minimum sample size estimated was 60 children (30 per group).

During implementation, however, a much larger number of eligible children were identified and recruited. Ultimately, 221 MAM children were enrolled, and 136 children completed the endpoint assessment (FMD = 71; BMD = 65). This achieved sample size was more than double the originally planned minimum, providing substantially greater statistical precision.

A post-hoc power analysis, using the observed standard deviation and the achieved final sample (n = 136), shows that the study had >95% power to detect the originally assumed effect size under the same alpha level of 0.016. Thus, the final realised sample size provided significantly higher power than originally planned, strengthening confidence in the robustness of the study’s findings.

### Randomization and Masking

A total of 2396 children aged 18-59 months were screened across 93 AWCs. Of these, 221 MAM children (WHZ –2 to –3 SD) were enrolled, and 136 completed the endpoint assessment, with no prior diagnosis or treatment for undernutrition. An additional 84 healthy children (WHZ –1 to +1 SD) with no history of malnutrition were included as a comparison group. Children with congenital or acquired disorders, severe anaemia (<8 mg/dL), recent infections or hospitalisations, or those receiving concurrent treatment were excluded. Written informed consent was obtained from parents or guardians

### Intervention

Two ready-to-use therapeutic food (RUTF) products: Finger Millet with Dates (FMD) and Balamrutham Plus with Dates (BMD), were developed for the intervention. FMD was formulated using the finger millet variety GPU-28, predominantly cultivated in Karnataka, Andhra Pradesh, and Tamil Nadu, India (ICAR-IIMR, 2023). Processing steps included 24-h soaking, 48-h germination, and roasting, which demonstrated optimal nutrient retention. The germinated–roasted flour was gamma irradiated (8 kGy) to ensure microbial safety. All raw ingredients were evaluated for microbial load and aflatoxin contamination following FSSAI SOP 11014/07/2021-QA, and results showed <10 cfu/g and no detectable aflatoxins.

BMD was prepared following the standard Balamrutham Plus formulation protocol (Figure 1), with vitamin–mineral fortification aligned to the existing product specifications. Both formulations provided >350 kcal per serving, with balanced proportions of protein, fat, fiber, and minerals. FMD was notably higher in insoluble fiber and micronutrient content.

**Figure 1:**
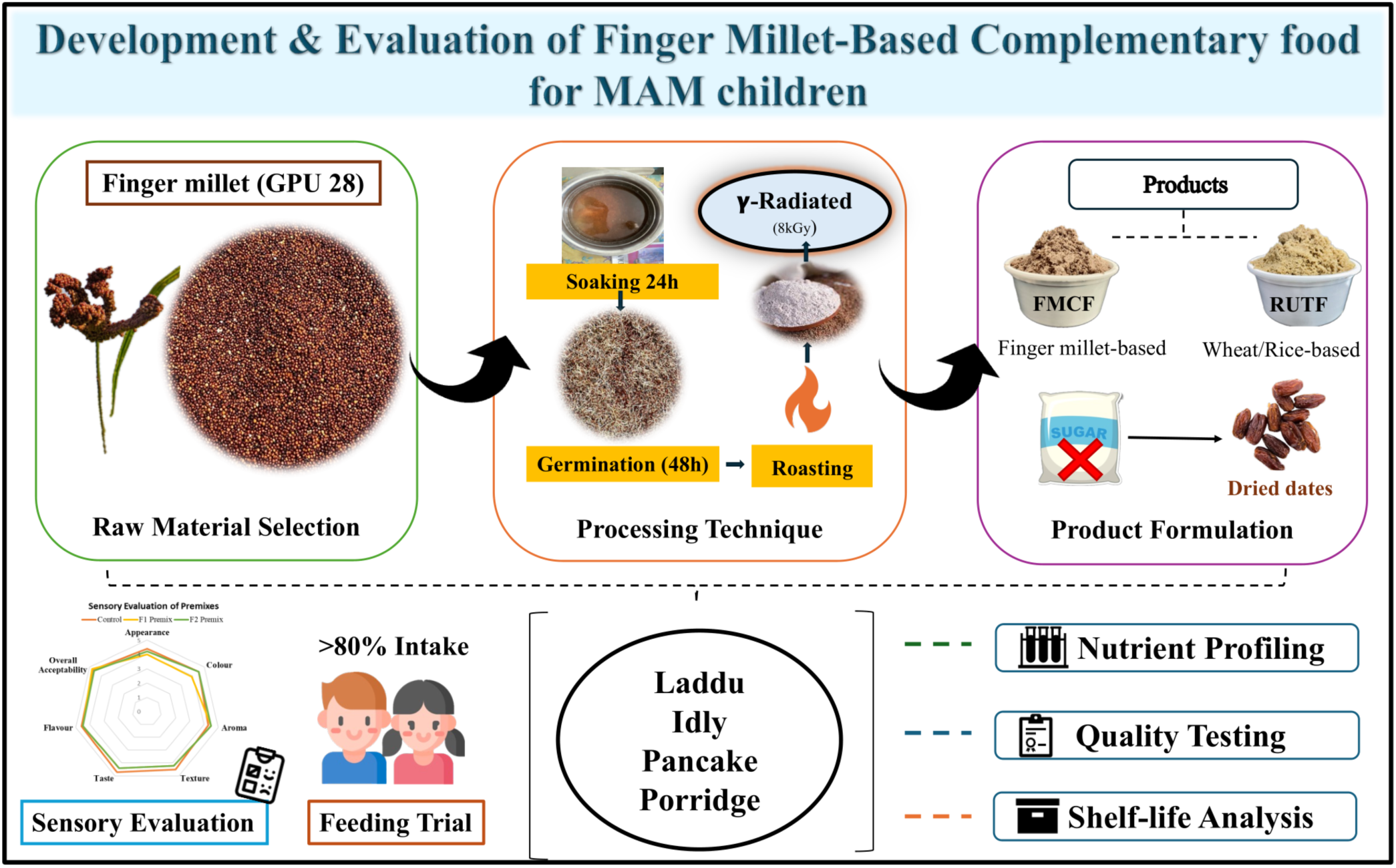
Overview of the Product development work flow.

Recipes made using FMD and BMD: porridge, laddu, idly, and pancake, were standardized to 30 g per serving. Sensory evaluation of FMD, BMD, their recipes, and the control (existing Balamrutham Plus) was conducted with 30 semi-trained and trained panelists (Anganwadi workers, mothers, staff, and students) using a 5-point hedonic scale. Data were analyzed using independent t-tests. FMD received a mean overall acceptability score of 4.8 (p > 0.05). Panelists particularly favored FMD recipes for flavour, texture, and aroma, with idly being the most preferred, followed by ladoo, pancake, and porridge (p < 0.05). In field acceptability trials across AWCs, children consumed >75% of each prepared recipe, indicating high acceptability.

MAM children were allocated to receive either FMD or BMD supplementation for 8 weeks. Anganwadi teachers and caretakers were trained in recipe preparation and feeding practices. Each AWC was provided with product packets per child, an idly cooker, pan, scoops, and a spatula. Caregivers were given food diaries and adverse event reporting forms to document daily intake and report any allergic reactions or digestive issues (constipation, diarrhoea), as well as symptoms such as fever, cough, vomiting, abdominal pain, bloating, or skin rashes throughout the intervention. Feeding sessions were closely supervised and routinely monitored to ensure compliance and safety. Further procedural details provided in our preprint article (Rai S et al., 2025).

### Nutrition education

The caregivers and mothers received nutrition counseling and education on sanitation and hygiene practices that should be followed regularly to ensure the well-being of their children and family members. They were clearly informed about managing undernutrition through a healthy diet, emphasizing increased protein intake from sources like eggs, milk, and legumes, as well as the importance of consuming adequate fruits and vegetables, green leafy vegetables, maintaining proper mealtime/frequency, limited intake of junk foods and staying hydrated to prevent severe consequences. They also learned about the significance of the study and the benefits of millet consumption. Individual counseling and group discussions were conducted to reinforce these messages.

### Food intervention and data collection

MAM children were designated to either FMD or BMD supplementation for a period of over 8 weeks. AW teachers/caretakers were trained for the recipe preparation and feeding. In each AWC, product packets per subject, idly cooker, pan, scoops, and spatula were distributed.

Food diary and adverse event forms were given to track daily consumption and report any allergic reactions or digestive issues, such as constipation or diarrhea, as well as symptoms like fever, cough, vomiting, abdominal pain, bloating, or skin rashes during the intervention. The feeding process was closely supervised and regularly monitored.

## Outcome Measures

### Anthropometry: *Weight, height, MUAC, Z-scores*

Children’s height, weight, and mid-upper arm circumference (MUAC) were measured at the Anganwadi Centers by trained field investigators. Weight was measured to the nearest 0.1 kg using a SECA 813 digital weighing scale, with participants standing barefoot and wearing minimal clothing. The weighing scale was calibrated daily using certified standard weights. Height was measured to the nearest 0.1 cm using a SECA 213 portable stadiometer, with children standing upright, heels together, and aligned to the measurement plane. MUAC was measured on the left upper arm at the midpoint between the acromion and olecranon using a non-stretch, fiber-reinforced MUAC tape.

Weight-for-height z-scores (WHZ) were computed according to WHO (2007) and Indian Academy of Pediatrics (IAP) 2015 growth standards using the Anthro Cal application (AIIMS, New Delhi).

## Body Composition

Body composition was assessed using the InBody S10, a portable, medical-grade, 4-compartment body composition analyzer based on Direct Segmental Multi-Frequency Bioelectrical Impedance Analysis (DSM-BIA). Parameters measured included total body water with segmental distribution, fat-free mass, body fat mass, body fat percentage, skeletal muscle mass, visceral fat area, basal metabolic rate (BMR), body cell mass (BCM), and bone mineral content (BMC).

Measurements were obtained using tetrapolar 8-point tactile electrodes attached to the thumbs and middle fingers of both hands and the ankle points of both feet. Assessments were conducted before mid-day, with participants in standing, seated, or supine posture, ensuring no contact with metal surfaces and maintaining a stable ambient temperature. All readings were recorded and analyzed according to the manufacturer’s guidelines.

## Dietary Recall

Dietary intake was assessed using a three-day 24-h dietary recall for each participant and nutrient values were derived from the Indian Food Composition Tables (IFCT, 2017). Macronutrient adequacy was evaluated using the Acceptable Macronutrient Distribution Range (AMDR), which recommends 50–60% of total energy from carbohydrates, 5–15% from protein, and 30–40% from fats for children aged 1–3 years, and 30–35% fat for children aged 4–5 years (ICMR–NIN Expert Group, 2020).

The probability of adequacy (PA) for seven micronutrients was estimated using the Estimated Average Requirements (EARs) set by the Institute of Medicine. Standard deviations of requirements were calculated using the coefficient of variation (CV) formula: SD = CV × EAR. A CV of 20% was applied for vitamin A, and 10% for all other micronutrients. Iron and zinc bioavailability were assumed at 5% for calculation of physiological requirements. The mean probability of adequacy (MPA) was computed as the average PA across all micronutrients. An MPA < 0.5 was considered indicative of inadequate micronutrient intake (Madhari et al., 2019).

## Statistical Analysis

All analyses were conducted using the intention-to-treat approach, with a supportive per-protocol analysis restricted to children who consumed at least 80% of their allocated supplement according to daily food-consumption logs. Data underwent quality checks that included verification against field registers, range checks for anthropometric indicators, and identification of outliers using ±3 SD criteria based on WHO reference standards. Derived variables such as WHZ, HAZ, percentage change from baseline, and micronutrient probability of adequacy were computed using WHO and FAO/INFOODS algorithms.

Baseline characteristics were summarised using means and standard deviations or medians and interquartile ranges for continuous variables, and proportions for categorical variables. Differences between the FMD and BMD groups at baseline were examined using t-tests or Mann-Whitney U tests for continuous variables and χ² or Fisher’s exact tests for categorical variables.

To evaluate changes in anthropometric outcomes (weight, height, MUAC) over time, repeated-measures linear mixed-effects models were applied, with fixed effects for group, time, and the group × time interaction, and a random intercept for each participant to account for within-child correlation. These models were used to compare trajectories at Days 20, 40, 100, and 160. Estimated marginal means with 95% confidence intervals were reported, along with absolute and percentage changes from baseline. For endpoint comparisons at Days 40 and 160, ANCOVA models adjusted for baseline values were used as supportive analyses.

Body-composition outcomes obtained from the InBody S10 (fat mass, fat-free mass, skeletal muscle mass, bone mineral mass) were analysed using ANCOVA models adjusted for baseline values. Standardised effect sizes (Cohen’s d) were calculated to aid interpretation of magnitude. Dietary intake from the three-day recall was analysed by computing mean energy and macronutrient intake and the proportion of children meeting AMDR recommendations; group comparisons were carried out using t-tests or χ² tests as appropriate. Micronutrient adequacy was quantified using the probability-of-adequacy method and mean probability of adequacy (MPA), and differences in PA and MPA between groups were assessed using t-tests or non-parametric equivalents.

Morbidity outcomes such as fever, diarrhoea, vomiting, and respiratory symptoms were summarised as episode counts and compared across groups using χ² tests. Relative risks with 95% confidence intervals were reported. Compliance was summarised as median percentage adherence and compared using the Wilcoxon rank-sum test; exploratory linear regression models were used to assess associations between compliance and anthropometric gains.

Subgroup analyses explored variations in effect by sex, age category (18-36 vs 36-59 months), baseline severity of wasting, and high versus low compliance. Interaction terms were tested within linear mixed-effects models to evaluate effect modification. Missing data assumed to be missing at random were handled using maximum-likelihood estimation inherent to mixed models, with multiple imputation used as a sensitivity analysis for key endpoints.

Because the final analysed sample (n = 136) exceeded the originally calculated requirement (n = 60), a post-hoc power calculation was performed using the observed mean differences and standard deviations. This analysis showed that the study had >95% power to detect the originally assumed effect sizes at an alpha level of 0.016. All statistical analyses were conducted using Stata 15 and R, and results were presented with two-sided p-values and 95% confidence intervals in accordance with CONSORT reporting standards.

## Ethical Approval

Ethical approval was obtained from the Institutional Ethics Committee (IEC No. CR/2/V/2023), and the trial was prospectively registered with the Clinical Trials Registry-India (CTRI/2023/06/053590).

## Results

### Participant flow

Of the 2396 children screened at Anganwadi centres, 460 met the study’s criteria for Moderate acute malnourished and 221 were recruited. Eligible participants were randomly assigned to either the finger millet-based dietary intervention (FMD, n=112) and the Balamrutham-based dietary intervention (BMD, n=109). An additional 1840 Healthy children screened separately, of whom 250 were recruited for comparison. Retention during the supervised intervention remained high. 73 from FMD whereas 66 from BMD groups completed the end point assessment. Minimal loss to follow-up was observed, with 71 (FMD) and 65 (BMD) participants completing the final evaluation. The flow diagram (Figure 2) illustrates clear progression from screening through randomisation, intervention, and follow-up, demonstrating balanced allocation and strong participant retention in both intervention arms.

**Figure 2.**
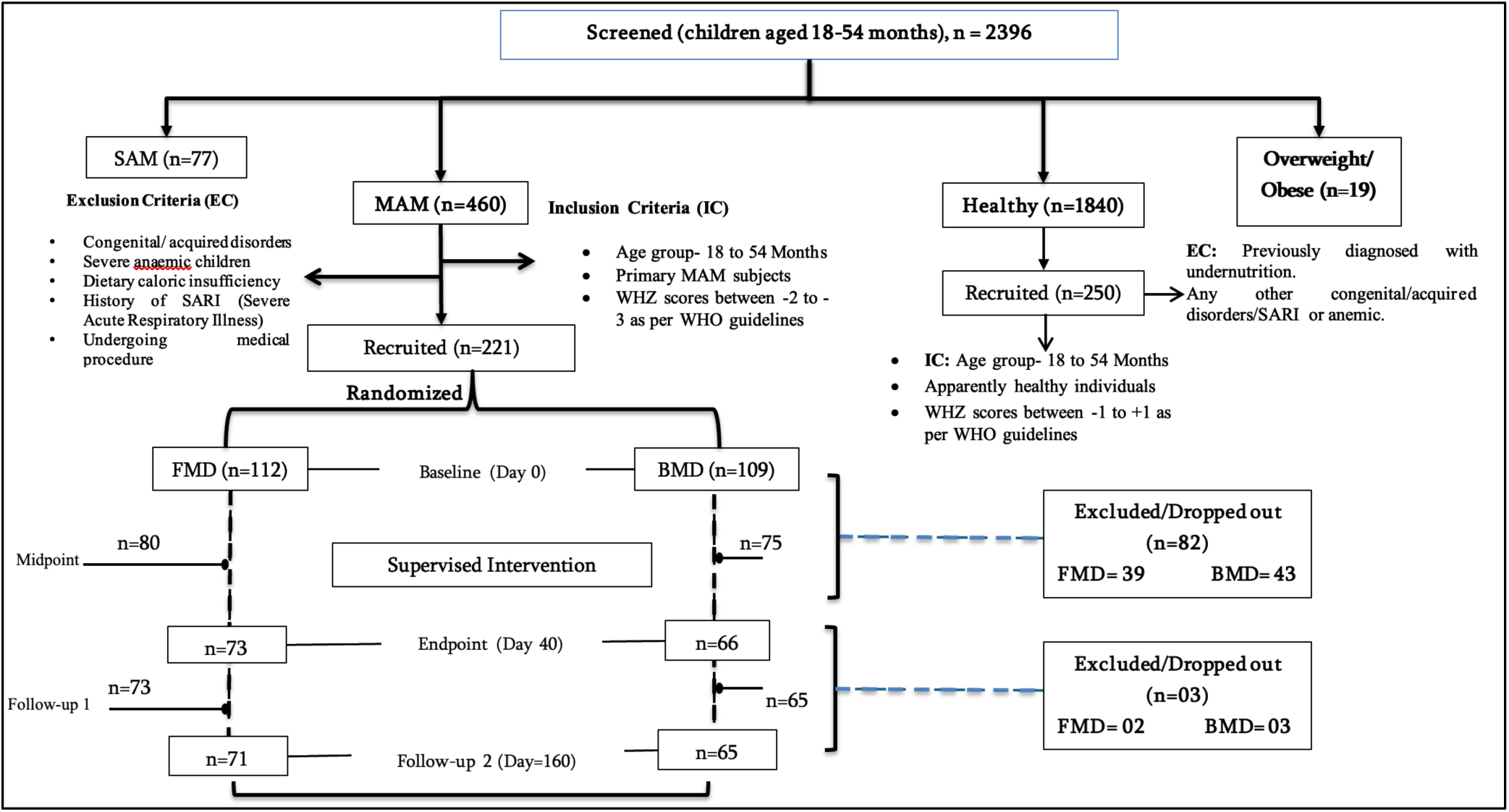
Study flow diagram for screening, enrolment, randomisation, intervention, and follow-up.

### Baseline characteristics

Demographic and dietary variables From 221 MAM subjects recruited, 136 completed the trial. Among them 71and 65 were given FMD and BMD respectively. Table 1 presents the demographic and baseline characteristics of the study population.

**Table 1.** Baseline characteristics of participants: demographic profile and dietary variables.

| Variables |  | Overall<br>(n= 136) | F1<br>(n= 71) | F2<br>(n= 65) | p-value |
| --- | --- | --- | --- | --- | --- |
| Sex, n (%) | Female | 53 (39.0) | 26 (40.0) | 27 (38.0) | 0.814 |
|  | Male | 83 (61.0) | 39 (60.0) | 44 (62.0) |  |
| Age in months, mean (SD) | - | 34.89<br>(9.38) | 33.93<br>(9.22) | 35.78<br>(9.50) | 0.252 |
| Birth Weight, mean (SD) | - | 2.73<br>(0.37) | 2.73<br>(0.38) | 2.73<br>(0.38) | 0.934 |
| Mode of Delivery, n (%) | Normal | 48 (42.1) | 25 (47.2) | 23 (37.7) | 0.345 |
|  | C-Section | 66 (57.9) | 28 (52.8) | 38 (62.3) |  |
| Birth Order, n (%) | First | 35 (30.7) | 11 (20.8) | 24 (39.3) | 0.100 |
|  | Second | 50 (43.9) | 29 (54.7) | 21 (34.4) |  |
|  | Last | 29 (25.4) | 13 (24.6) | 16 (26.3) |  |
| Diet preference of the child, n (%) | Veg | 2 (1.8) | 0 (0.0) | 2 (3.3) | 0.366 |
|  | Non-Veg | 109 (95.6) | 52 (98.1) | 57 (93.4) |  |
|  | Veg + Egg | 3 (2.6) | 1 (1.9) | 2 (3.3) |  |
| Mother's Education, n (%) | Illiterate | 11 (9.7) | 7 (13.2) | 4 (5.5) | 0.416 |
|  | Primary Education | 23 (20.2) | 12 (22.6) | 11 (18.0) |  |
|  | Secondary Education | 26 (22.8) | 11 (20.8) | 15 (24.6) |  |
|  | Higher | 54 (47.34) | 23(43.5) | 31 (50.8) |  |
| **Father's Education, n (%)** | Illiterate | 10 (8.8) | 8 (15.1) | 2 (3.3) | 0.144 |
|  | Primary Education | 27 (23.7) | 14 (26.4) | 13 (21.3) |  |
|  | Secondary Education | 36 (31.6) | 13 (24.5) | 23 (37.7) |  |
|  | Higher | 41 (36.0) | 18 (34.0) | 23(37.8) |  |

### Demographic details

Among the participants, 39% were female and 61% were male with a similar distribution between the BMD and FMD groups (p=0.814). The mean age of the participants was 34.89 months with no significant difference between groups (p=0.252). The mean birth weight was consistent across groups at 2.73kg (p=0.934).

### Birth and Family Characteristics

Regarding mode of delivery, 42.1% of the children were born through normal delivery, while 57.9% were delivered via C-section, with no significant difference between groups (p=0.345). Majority of children were either first or second born, with birth order distributions not differing significantly (p=0.100).

### Religious and Social backgrounds

Most participants identified as Hindu (84.6%), followed by Muslim (9.4%) and Christian (6.0%) with similar distributions between groups (p=0.967). The caste distribution showed that 56.1% belonged to Other Backward Caste followed by Scheduled Caste (30.7%), General (9.7%) and Scheduled Tribes (3.5%) though differences were not statistically significant (p=0.302).

### Dietary preferences

Majority of children (95.6%) consumed a non-vegetarian diet, while a small percentage followed a vegetarian or eggetarian diet. There was no considerable difference in dietary habits between groups (p=0.366).

### Parental characteristics

The mean age of mothers was 27.54 years and fathers had a mean age of 32.36 years with no significant variation. Educational levels varied with 47.34% of mothers and 36% of fathers having attained higher education. While differences in parental education status were observed, they did not reach statistical significance.

Overall, no statistically significant differences were observed between FMD and BMD groups across the assessed baseline characteristics.

### Food Intervention data analysis

Children in both groups demonstrated high compliance, consuming the study products on over 80% of intervention days, with an average intake of 120 g/day. Consumption frequency was comparable between the FMD and BMD groups, indicating similar adherence patterns and good acceptability of both products. Figure 3 summarises these compliance indicators using box plots of food-consumption percentage and total consumption days. While BMD showed a slightly higher median consumption percentage than FMD (p = 0.048), the number of consumption days did not differ significantly (p = 0.484). Overall, the findings confirm strong and consistent intake across both intervention arms.

**Figure 3.**
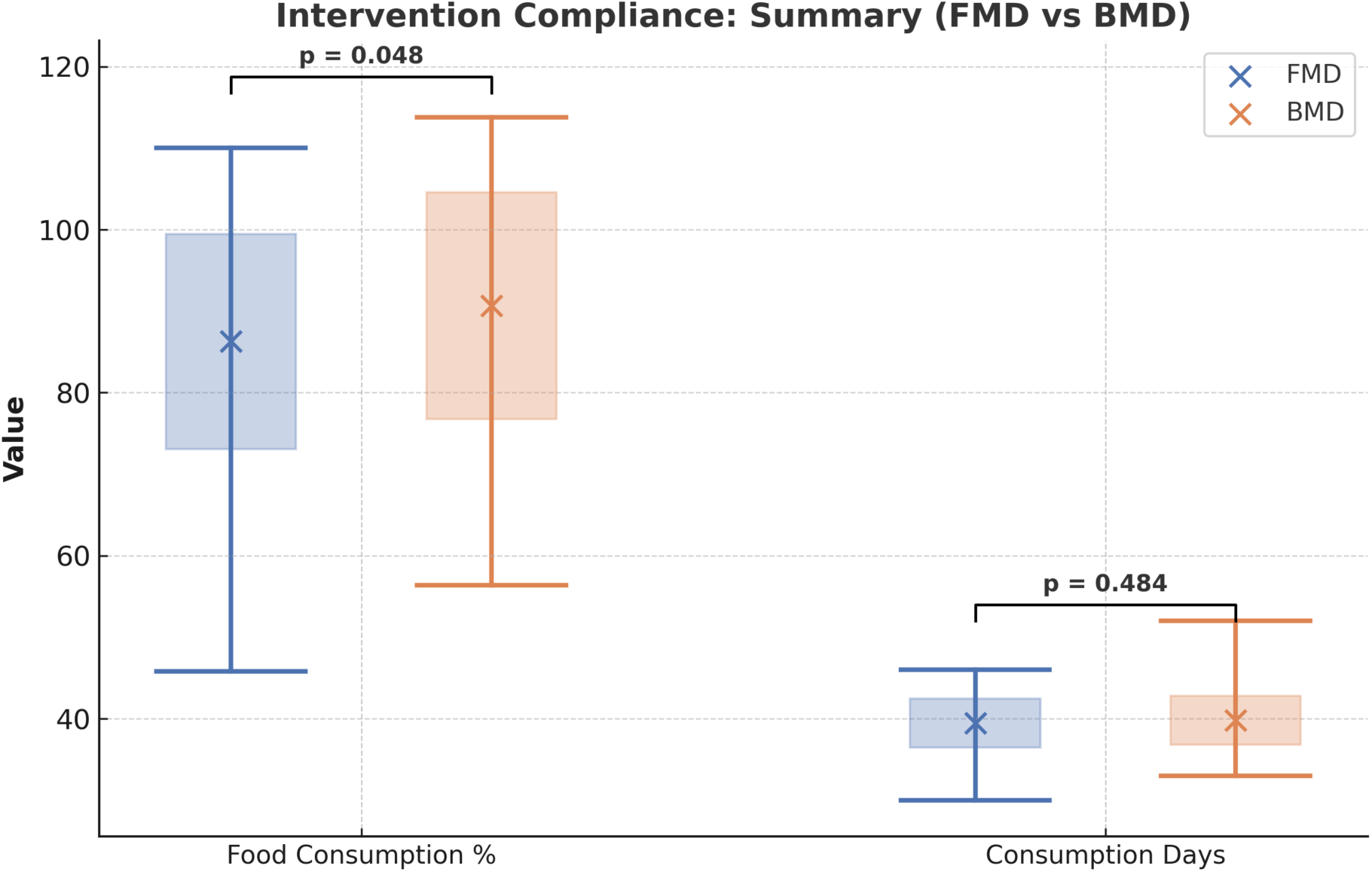
Intervention compliance based on food consumption proportion and number of consumption days among FMD and BMD groups.

### Primary outcomes: Anthropometric gains

At the outset, growth indicators such as weight, height, and MUAC were comparable between the two groups. Across the 40-day intervention, both supplements supported steady improvements. Weight increased by 4.6-4.7% by day 20 and 6.3-6.4% by day 40, while height rose by 1.6% at day 20 and approximately 2.4% at day 40. MUAC showed similar upward trends, increasing by 1.92% (BMD) and 4.13% (FMD) at day 20, and by 3.46% and 6.19%, respectively, at day 40. Confidence intervals consistently overlapped across all parameters, indicating comparable effects of both interventions during the active supplementation phase (Table 2a). Growth continued during the post-intervention follow-up period even after supplementation ceased. By day 100, weight gain reached 7.31-7.44%, height increased to 3.69-3.77%, and MUAC rose to 4.40-5.42%. By day 160, these improvements further reached 9.83-10.01% for weight, 5.03-5.19% for height, and 4.60-6.74% for MUAC. Final values for all three anthropometric measures remained closely aligned between the groups, with no differential effect observed at any timepoint. Table 2b showed that children receiving FMD experienced a significantly greater improvement in WHZ at endline compared with those on the cereal-based product (BMD), indicating better short-term recovery from wasting. Improvements in WAZ and HAZ were observed in both groups, with no significant between-group differences.

**Table 2a.**
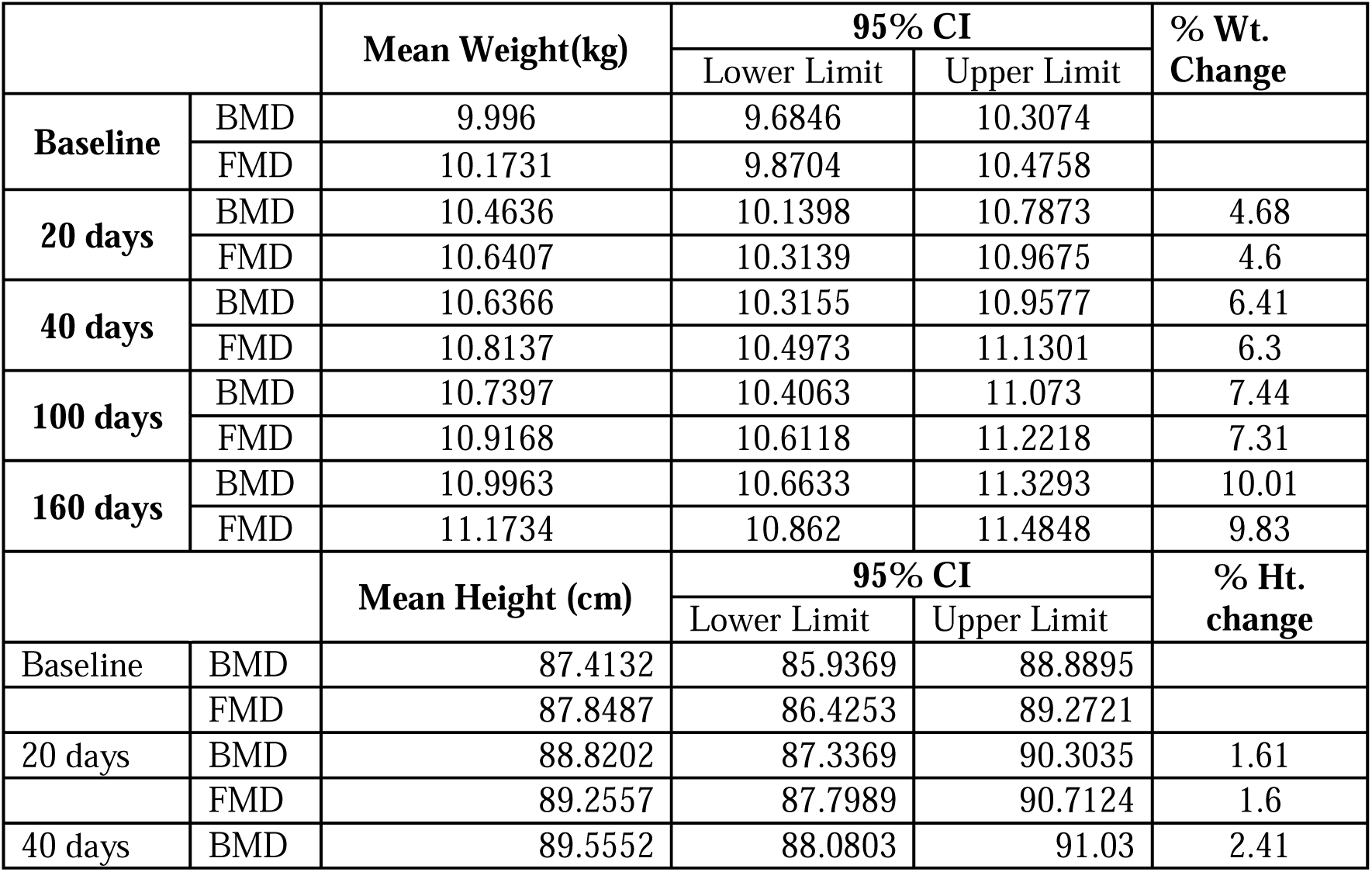
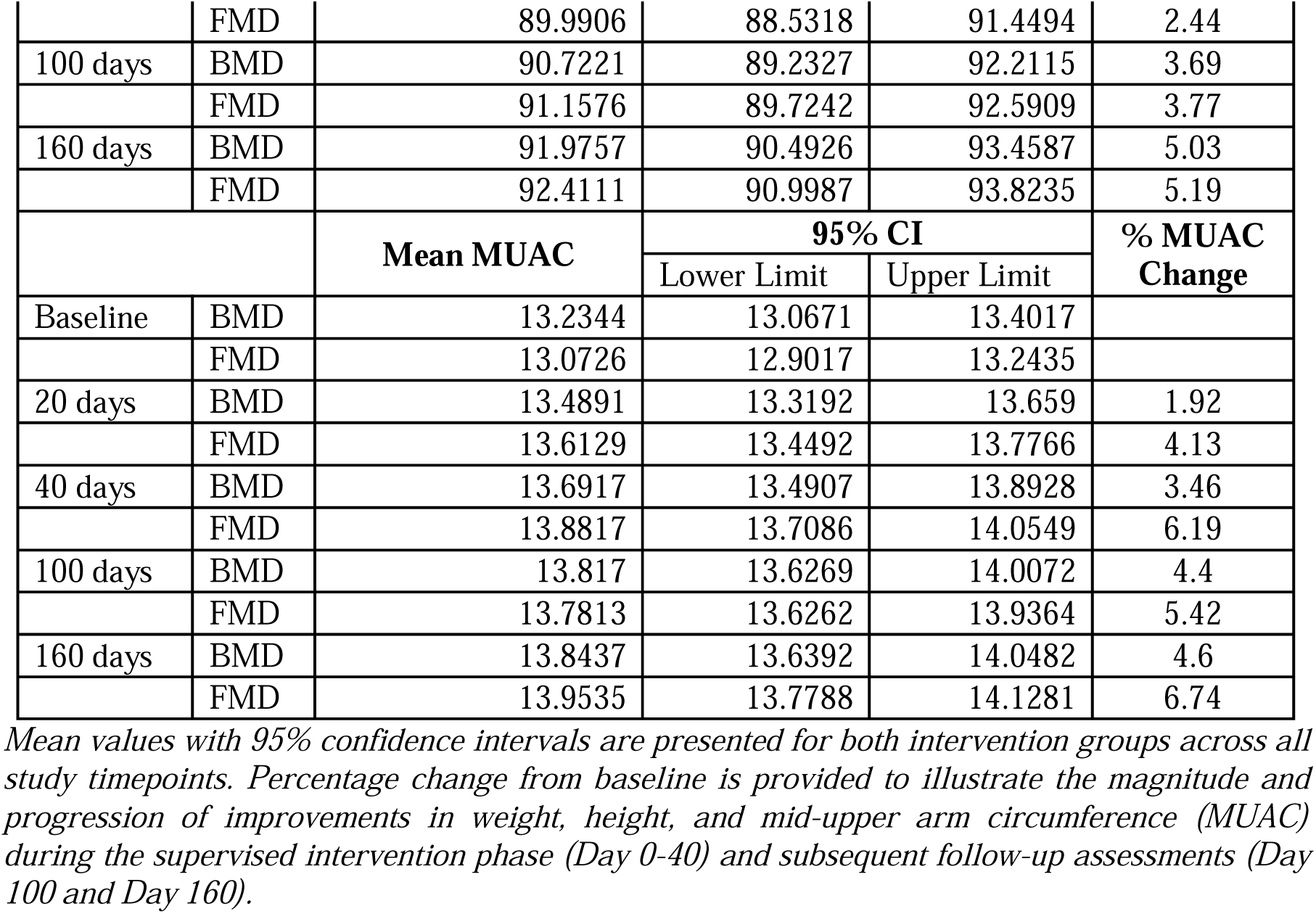
Anthropometric outcomes (weight, height, and MUAC) during the intervention and follow-up period among children with moderate acute malnutrition.

|  |  | Mean Weight(kg) | 95% CI |  | % Wt. Change |
| --- | --- | --- | --- | --- | --- |
|  |  |  | Lower Limit | Upper Limit |  |
| Baseline | BMD | 9.996 | 9.6846 | 10.3074 |  |
|  | FMD | 10.1731 | 9.8704 | 10.4758 |  |
| 20 days | BMD | 10.4636 | 10.1398 | 10.7873 | 4.68 |
|  | FMD | 10.6407 | 10.3139 | 10.9675 | 4.6 |
| 40 days | BMD | 10.6366 | 10.3155 | 10.9577 | 6.41 |
|  | FMD | 10.8137 | 10.4973 | 11.1301 | 6.3 |
| 100 days | BMD | 10.7397 | 10.4063 | 11.073 | 7.44 |
|  | FMD | 10.9168 | 10.6118 | 11.2218 | 7.31 |
| 160 days | BMD | 10.9963 | 10.6633 | 11.3293 | 10.01 |
|  | FMD | 11.1734 | 10.862 | 11.4848 | 9.83 |
|  |  | Mean Height (cm) | 95% CI |  | % Ht. change |
|  |  |  | Lower Limit | Upper Limit |  |
| Baseline | BMD | 87.4132 | 85.9369 | 88.8895 |  |
|  | FMD | 87.8487 | 86.4253 | 89.2721 |  |
| 20 days | BMD | 88.8202 | 87.3369 | 90.3035 | 1.61 |
|  | FMD | 89.2557 | 87.7989 | 90.7124 | 1.6 |
| 40 days | BMD | 89.5552 | 88.0803 | 91.03 | 2.41 |
|  | FMD | 89.9906 | 88.5318 | 91.4494 | 2.44 |
| 100 days | BMD | 90.7221 | 89.2327 | 92.2115 | 3.69 |
|  | FMD | 91.1576 | 89.7242 | 92.5909 | 3.77 |
| 160 days | BMD | 91.9757 | 90.4926 | 93.4587 | 5.03 |
|  | FMD | 92.4111 | 90.9987 | 93.8235 | 5.19 |
|  |  | Mean MUAC | 95% CI |  | % MUAC Change |
|  |  |  | Lower Limit | Upper Limit |  |
| Baseline | BMD | 13.2344 | 13.0671 | 13.4017 |  |
|  | FMD | 13.0726 | 12.9017 | 13.2435 |  |
| 20 days | BMD | 13.4891 | 13.3192 | 13.659 | 1.92 |
|  | FMD | 13.6129 | 13.4492 | 13.7766 | 4.13 |
| 40 days | BMD | 13.6917 | 13.4907 | 13.8928 | 3.46 |
|  | FMD | 13.8817 | 13.7086 | 14.0549 | 6.19 |
| 100 days | BMD | 13.817 | 13.6269 | 14.0072 | 4.4 |
|  | FMD | 13.7813 | 13.6262 | 13.9364 | 5.42 |
| 160 days | BMD | 13.8437 | 13.6392 | 14.0482 | 4.6 |
|  | FMD | 13.9535 | 13.7788 | 14.1281 | 6.74 |

Mean values with 95% confidence intervals are presented for both intervention groups across all study timepoints. Percentage change from baseline is provided to illustrate the magnitude and progression of improvements in weight, height, and mid-upper arm circumference (MUAC) during the supervised intervention phase (Day 0-40) and subsequent follow-up assessments (Day 100 and Day 160).

**Table 2b:** Anthropometric z-scores comparison between FMD vs BMD across time points.

| Mean ± SD | Timepoint | FMD (n=71) | BMD (n=65) | P value |
| --- | --- | --- | --- | --- |
| **WHZ** | Base | -2.35 ± 0.31 | -2.37 ± 0.29 | 0.772 |
| | End | -2.00 ± 0.75 | -2.24 ± 0.57 | 0.042 |
| | Follow up | -2.23 ± 0.56 | -2.35 ± 0.57 | 0.233 |
| **WAZ** | Base | -2.64 ± 0.62 | -2.54 ± 0.66 | 0.394 |
| | End | -2.06 ± 0.76 | -2.09 ± 0.73 | 0.791 |
| | Follow up | -1.80 ± 0.74 | -1.79 ± 0.77 | 0.976 |
| **HAZ** | Base | -1.87 ± 1.05 | -1.69 ± 1.15 | 0.352 |
| | End | -1.27 ± 1.14 | -1.06 ± 1.19 | 0.309 |
| | Follow up | -0.55 ± 1.24 | -0.41 ± 1.26 | 0.526 |

## Secondary outcomes

### Body composition changes

Both groups showed notable improvements in hydration and lean tissue indices, but FMD group demonstrated consistently higher gains across several parameters, including intracellular water (+8.27%), extracellular water (+8.03%), total body water (+8.02%),

protein mass (+8.99%), and fat-free mass (+9.62%). Children receiving FMD also showed greater increase in soft lean mass (+8.25%) and skeletal muscle mass (+19.11%) from baseline to endline, indicating enhanced lean tissue accretion. Percent body fat decreased in both groups, with a stronger reduction in the FMD group (–3.37%), reflecting a healthier shift toward metabolically active tissue. Mineral mass increased substantially in FMD (+42.86%), supporting improvements in structural tissue composition. Although between-group differences were not statistically significant. Figures 4a, 4b and 4c illustrated the individual mean scores and patterns of body composition across timepoints.

**Figure 4a:**
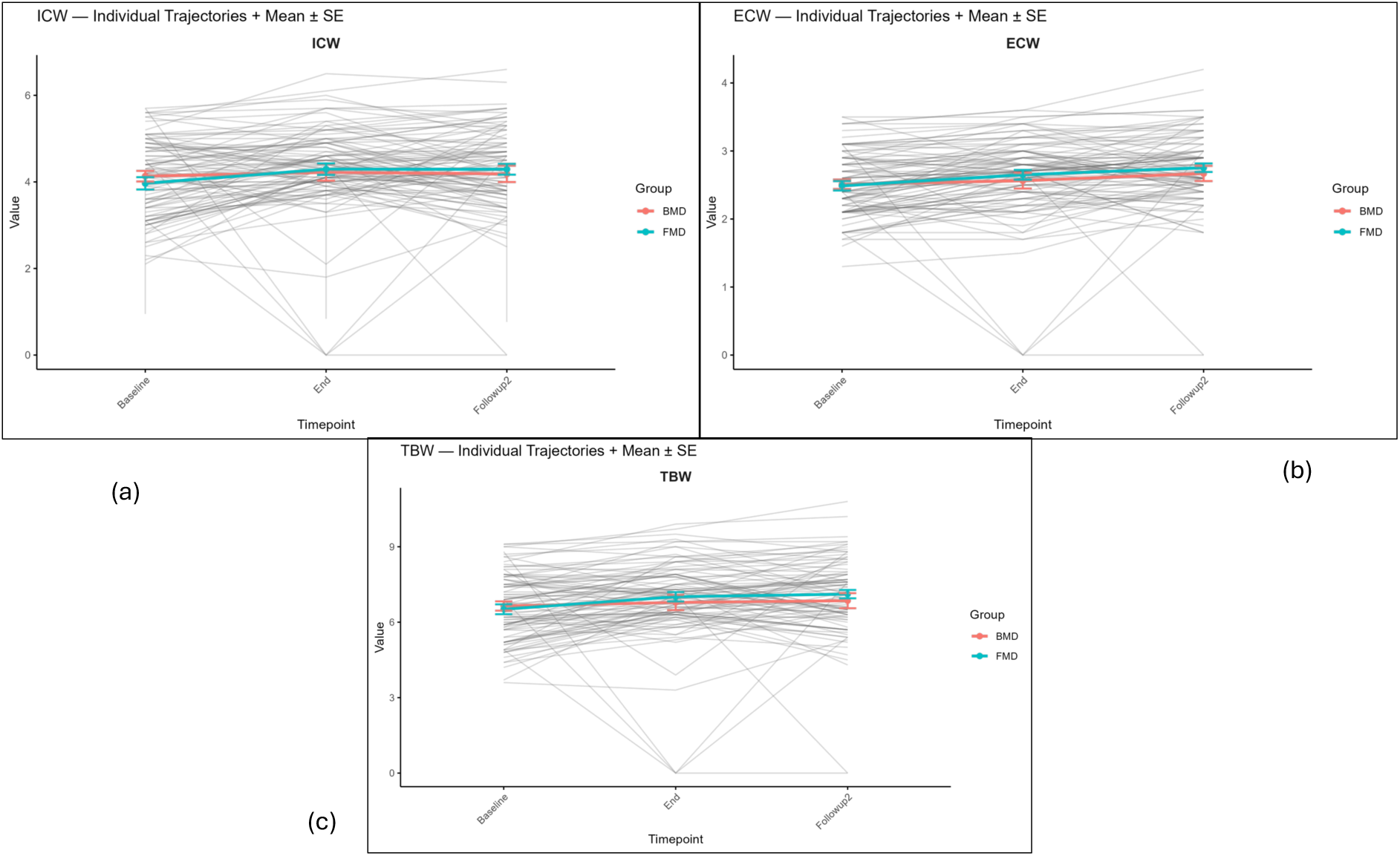
Plots depicting the individual mean ± SE and growth trajectories of Hydration indices (a) Intercellular water (ICW), (b) Extracellular water (ECW) and (c) Total body water (TBW).

**Figure 4b:**
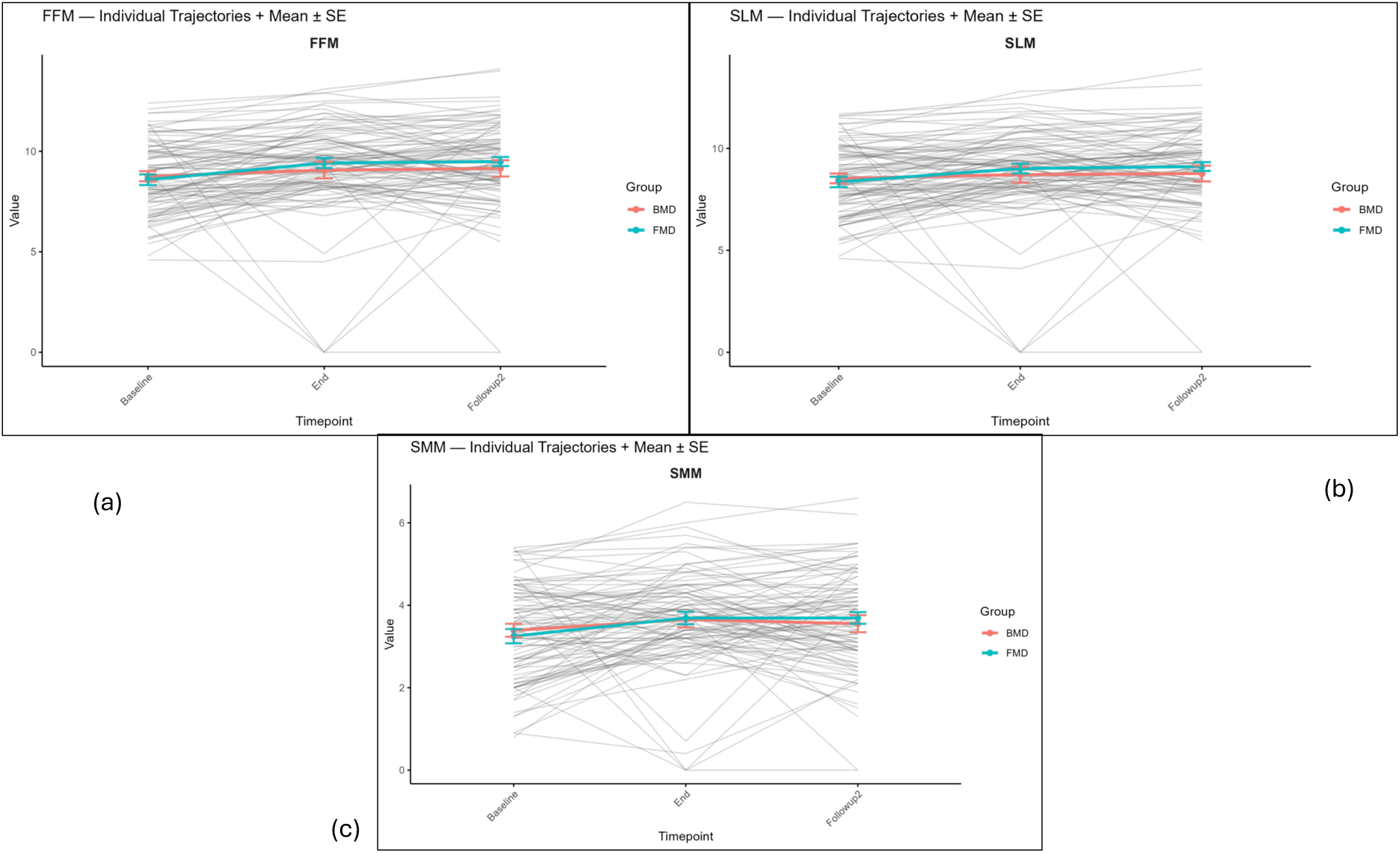
Plots depicting the individual mean ± SE and growth trajectories of Lean tissue mass indices (a) Fat free mass (FFM), (b) Soft lean mass (SLM) and (c) Skeletal muscle mass (SMM).

**Figure 4c:**
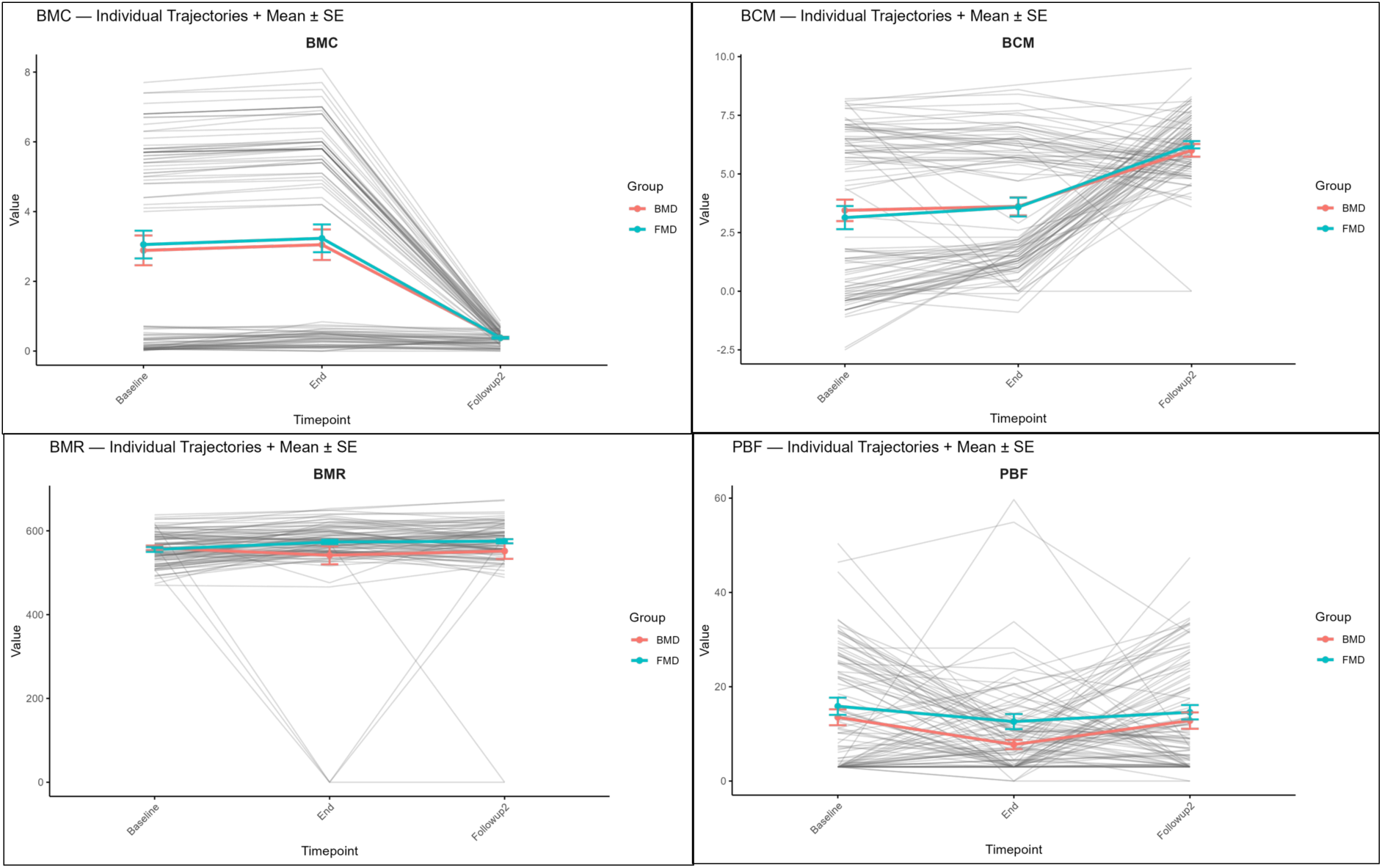
Plots depicting the individual mean ± SE and trajectories of Bone mineral content (BMC), Body cell mass (BCM), Basal metabolic rate (BMR) and Percent Body fat (PBF).

### Nutrient adequacy

Macronutrient distribution differed significantly between groups (Table 5). Healthy children had a higher proportion of energy from carbohydrates and fat (p<0.05), with more meeting AMDR recommendations compared with the FMD and BMD groups. Protein contribution to energy intake was similar across groups, with fewer than half satisfying the AMDR.

**Table 5.**
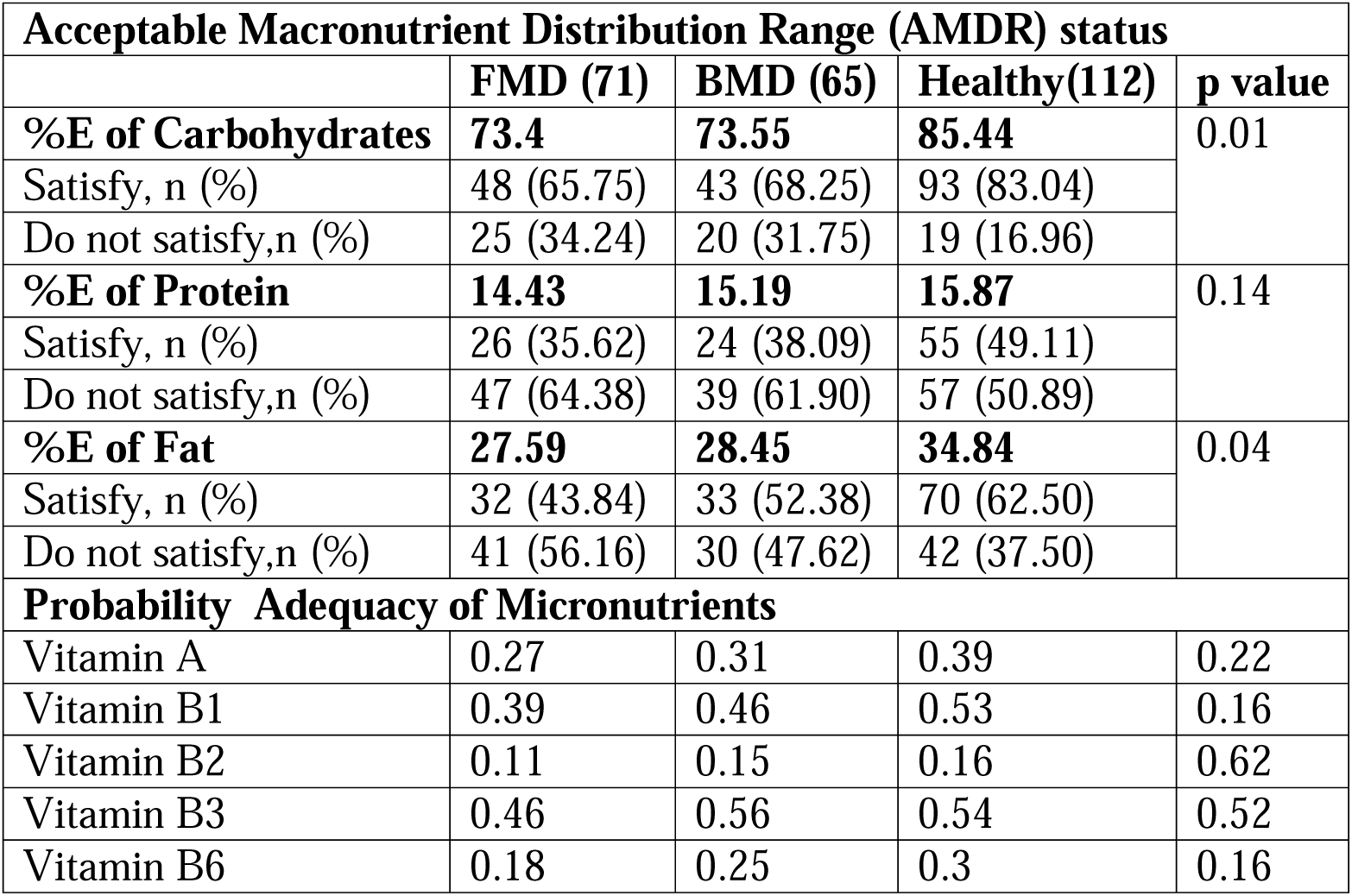
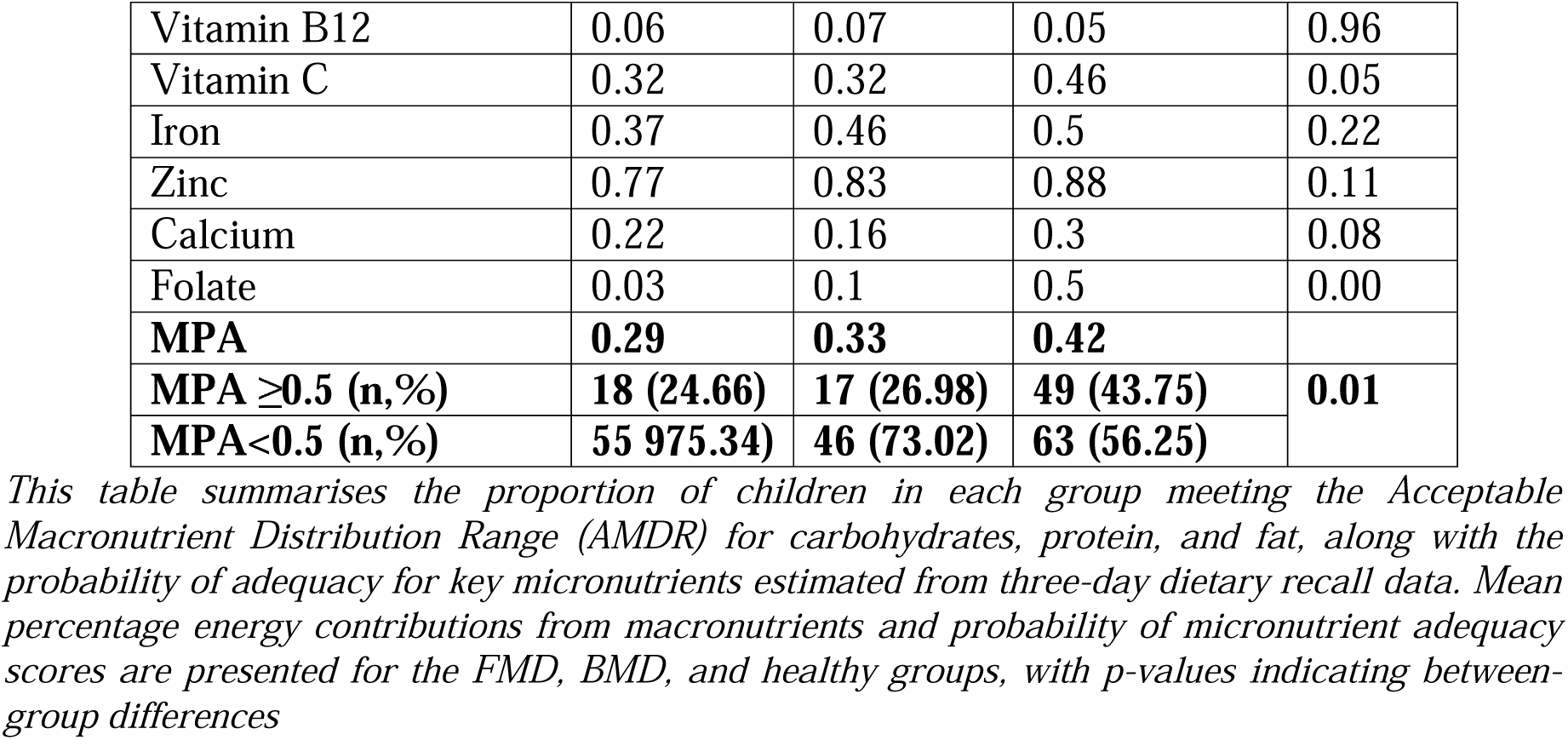
Acceptable Macronutrient Distribution Range (AMDR) status and probability of micronutrient adequacy based on three-day dietary recall among FMD, BMD, and healthy children.

Micronutrient adequacy was consistently lower among children with MAM than among healthy peers. Adequacy probabilities for vitamins A, C, B-complex, calcium, zinc, and especially folate were notably poorer in the FMD and BMD groups. Mean Probability of Adequacy (MPA) was highest in healthy children (0.42), and a significantly larger proportion achieved MPA ≥0.5 compared with intervention groups (p=0.01).

Overall, dietary quality among MAM children was markedly inferior to that of healthy children, with substantial deficiencies in both macro– and micronutrient adequacy.

### Other clinical outcomes

During the intervention, subjects receiving BMD and FMD reported a lower prevalence of diarrhea and vomiting (1 [3%] vs 0 [0%] and marginally lower rate incidence of fever, cough and cold cases (11 [36%] vs 19 [51%]) respectively. No food allergies or intolerance was reported.

## Discussion

This randomized controlled trial demonstrated that finger millet-based supplementation produced meaningful improvements in growth indicators among moderately acute malnourished (MAM) preschool children, with gains in weight, height, MUAC, and WHZ similar to those observed with the existing cereal-based RUTF. Growth trajectories remained steady beyond the active intervention, indicating sustained benefit. These findings reinforce the potential of millet-based complementary foods as a culturally acceptable and nutritionally robust alternative to cereal-based formulations currently used in national programs.

Our results align with evidence from earlier supplementation studies. Azmi et al. (2020) reported significant improvements in anthropometry after eight weeks of lipid-based supplementation in children aged 35-39 months. Similarly, Ayuni et al. (2023) demonstrated substantial increases in weight (32%) and height (68%) among severely malnourished children receiving nutrient-dense supplements. The magnitude of weight gain in our study (∼9-10%) closely approximates the effect sizes reported in prior trials involving fortified RUTFs and cereal-legume blends. Historical evidence from Devdas et al. (1984) also supports the advantage of finger millet over rice-based diets in improving weight, height, MUAC, and haemoglobin among preschoolers. A recent meta-analysis showing 25% improvement in weight, 39% in MUAC, and 28% in height following finger millet consumption resonates with the growth patterns observed in the present trial. The physiological basis for these improvements may relate to the distinct nutrient profile of finger millet. The high calcium content (∼364 mg/100 g), along with bioavailable magnesium and phosphorus, may contribute to the observed increases in skeletal muscle and bone mineral content. Improved mineral mass and bone indices in the finger millet group are consistent with findings by Anitha et al. (2021), who noted superior bone health and mineral retention with millet-based diets. Additionally, finger millet provides a balanced amino acid profile with relatively high digestibility when processed through germination and roasting. These steps reduce antinutritional factors such as phytates, tannins, and saponins, thereby enhancing mineral bioavailability and protein utilization. The rich insoluble fibre (cellulose, hemicellulose, lignin) and soluble prebiotic fibres (β-glucan, arabinoxylan) may modulate the gut microbiota, improving mucosal integrity and nutrient absorption, a mechanism previously highlighted in gut–growth research among malnourished children.

In comparison to cereal-based RUTF/RUSF used in multiple global settings, the finger millet-based formulation performed equivalently in growth recovery while offering additional benefits related to micronutrient density and gastrointestinal tolerance. The BMD group, designed to reflect the current ICDS and Telangana Balamrutham formulation, showed improvements due to its higher protein and fat content. However, the greater enhancement in mineral mass, bone mineral content, and skeletal muscle mass in the finger millet group points to potential advantages of millet-based diets over wheat-or rice-based formulations, particularly in regions with high micronutrient deficiencies. The intervention demonstrated high acceptability, reflected in >80% consumption rates and consistent intake across the trial. The inclusion of date powder instead of refined sugar notably improved palatability while contributing fibre, iron, and potassium aligning with emerging recommendations for natural sweeteners in paediatric complementary foods. Nutrition counselling for caregivers and Anganwadi workers likely contributed to strong adherence, underscoring the importance of integrated behaviour-change strategies alongside supplementation. The findings have important implications for national programs such as ICDS and POSHAN 2.0. Millets, particularly finger millet, are locally available, climate-resilient, culturally accepted, and aligned with the government’s mandate to integrate traditional grains into complementary feeding. Transitioning a portion of cereal-based RUTF/RUSF formulations toward millet-based alternatives could enhance dietary diversity, improve micronutrient adequacy, and reduce reliance on refined ingredients. Given the high acceptability and favourable growth outcomes observed in this trial, finger millet–based supplements could be scaled within Anganwadi centres as part of routine MAM management.

In summary, this study adds to the growing evidence supporting millets as an effective, sustainable, and policy-relevant strategy for improving child nutritional outcomes. Integrating finger millet-based products into national nutrition schemes may help accelerate India’s progress toward reducing childhood undernutrition and achieving POSHAN 2.0 targets.

## Conclusion

Supplementing with finger millet and date powder resulted in improvements compared to the current product in children with moderate acute malnutrition aged 18-59 months. Overall, the results were on par with the existing intervention. In short, our manuscript highlights how finger millet can help address undernutrition by being an affordable, nutritious option that should be more widely used in food programs.

## CRediT authorship contribution statement

Authorship contributions: **Nikhita B R**: Investigation, Methodology, Writing original draft; **Sakshi Rai**: Investigation, Writing; **Sahil Bipin Kumar Suthar**: Writing-Review and editing; **Parth Sarin**: Writing-Review and editing, software, data curation; **Aruna V. Reddy**: Data collection, curation and Formal analysis; **Vijayalakshmi G**: Data collection, curation and Formal analysis; **Hemant Mahajan**: Formal analysis, Software, Supervision, Validation, Visualization, Writing-Review and editing; **Santosh Kumar Banjara**: Supervision, Validation, Visualization, Writing-Review and editing; **Ananthan Rajendran**: Conceptualization, Methodology, Supervision, Validation, Visualization, Writing-Review and editing; **Naveen Kumar R**: Methodology, Supervision, Validation, Visualization, Writing-Review and editing; **Sourav Sengupta**: Conceptualization, Funding acquisition, Investigation, Project administration, Supervision, Visualization, Writing-Review and editing; **Devaraj J. Parasannanavar**: Conceptualization, Funding acquisition, Investigation, Project administration, Supervision, Visualization, Writing-Review and editing.

## Funding

This study has been funded by ICMR-Indian Council of Medical Research

## Informed Consent Statement

Informed consent was obtained from the parents/guardians of all the participants involved in the study. The consent form was provided in two languages: one local and one formal (Telugu and English).

## Data Availability Statement

The data supporting this study’s findings will be made available on reasonable request.

## Disclosure

The authors state that there are no financial or personal relationships that could have influenced the work presented in this paper

## Confidentiality and Dissemination policy

Participant data will be kept confidential and securely stored, accessible only to authorized personnel. Identifiable information will be coded to protect privacy. Results will be shared through publications and presentations without revealing participant identities, following ethical and regulatory standards.

## Declaration of Interests

The authors declare no competing interests.

## Data Sharing Statement

De-identified participant data and code will be available on reasonable request after publication.

## Notes

### Competing Interest Statement

The authors have declared no competing interest.

### Clinical Trial

CTRI/2023/06/053590

### Author Declarations

Ethical approval was obtained from the ICMR National Institute of Nutrition ethics committee (IEC No. CR/2/V/2023)

